# Predictive analytics that reflect disease burden – the cumulative COMET score

**DOI:** 10.1101/2022.06.03.22275909

**Authors:** Oliver Monfredi, Robert T Andris, Douglas E Lake, J Randall Moorman

**Affiliations:** Department of Internal Medicine, University of Virginia, Charlottesville VA USA; Center for Advanced Medical Analytics, University of Virginia, Charlottesville VA USA

**Author notes:** Competing interests: DEL and JRM own stock in Medical Predictive Science Corporation, Charlottesville, VA; JRM is a consultant for Nihon Kohden Digital Health Solutions, Irvine, CA.

## Abstract

Predictive analytics tools variably take into account data from the electronic medical record, lab tests, nursing charted vital signs and continuous cardiorespiratory monitoring data to deliver an instantaneous score that indicates patient risk or instability. Few, if any, of these tools reflect the risk to a patient accumulated over the course of an entire hospital stay. This approach fails to best utilize all of the collated data regarding the risk or instability sustained by the patient, and hence fails to fully characterize this to optimize the ability of treating clinicians to maximize the chances of a favorable outcome. We have built on our instantaneous CoMET predictive analytics score to generate the cumulative CoMET score (cCOMET), which sums all of the instantaneous CoMET scores throughout a hospital admission relative to a baseline expected risk unique to that patient. We have shown that higher cCOMET scores predict mortality, but not length of stay, and that higher baseline CoMET scores predict higher cCoMET scores at discharge/death. cCoMET scores were higher in males in our cohort, and added information to the final CoMET when it came to the prediction of death. In summary, if one is going to go to the trouble and expense of performing repeated measures when performing predictive analytics calculations, we have shown that including all of these measures in a cumulative way adds data to instantaneous predictive analytics, and could improve the ability of clinicians to predict deterioration, and improve patient outcomes in so doing.

## Introduction

Early detection of clinical deterioration in hospitalized patients allowing early medical intervention to mitigate or prevent this has benefits in terms of morbidity, mortality and healthcare economy.^1-4^ Utilizing the ‘HeRO’ score based on heart rate characteristics, our group has demonstrated, in the setting of a randomized controlled trial, that using only data from the ECG, an instantaneous risk score can be calculated that predicts the future onset of sepsis in very low birthweight infants, and allows clinicians to intervene earlier, decreasing mortality in this most vulnerable group of patients.^5^ The score is predicated on the finding of a signature of illness^6^ in the continuous cardiorespiratory monitoring data that can be detected using mathematical time-series analyses and mapped to the risk of an event in the next time window.

The ethos underlying the HeRO score was translated to the discovery of signatures of illness in adult^7-9^ and pediatric ICU^10^ patients, and adult ward patients.^8^ In these settings, risk is displayed as the ‘CoMET’ (‘<u>Co</u>ntinuous <u>M</u>onitoring of <u>E</u>vent <u>T</u>rajectories’) score. In addition to parameters extracted from continuous cardiorespiratory monitoring data, the score takes account of vital signs and lab results from the electronic health record (EHR).

The outputs that generate CoMET, which are the products of logistic regression, are instantaneous metrics of ‘cardiorespiratory’ and ‘cardiovascular instability’, and the output display charts the trajectory of this instability over the prior 3 hours to allow identification of patients with a deteriorating trajectory.^11-14^

A potential shortcoming of this however is that, while excellent at reflecting patient trajectory *now*, i.e. based on the prior 3 hours of data, the CoMET score is unable to reflect the cumulative burden of illness sustained by the patient throughout their hospital stay. As such, the score may fail to reflect all that we have learned about a given patient and their illness to date. We hypothesize that it would be better to develop a predictive analytics tool that is reflective of all accrued data since the patient’s admission, to better refine our ability to predict outcomes and forewarn of deterioration.

Herein, we investigate the value of the novel cumulative CoMET (‘cCoMET’) score, which represents a continuous summation of the CoMET scores throughout a patient’s hospital stay, which we hypothesize will reflect the accumulating burden of physiological instability, and in doing so will be related to morbidity and mortality. This particular metric is akin to the HbA1C measured in diabetes, and is a ‘test with memory’, so that regardless of whether there is little or much *instantaneous* cardiorespiratory instability, the events that have gone before will still be reflected in the output predictive metric. We have previously explored this concept using the cumulative HeRO score in very low birth weight infants, demonstrating that a high cumulative HeRO score is significantly associated with in-hospital mortality.^6^ We hypothesize that the cCOMET will have the ability to summarize the entire hospital stay to date, and thereby give a more accurate and complete idea of the physiological insult suffered by a patient, which should assist in accurately predicting their trajectory and ultimately their outcome, and assist clinicians to direct their efforts to the most vulnerable and fragile patients under their care.

## Methods

### The Study Population

We studied adult (age =/> 18 years) patients consecutively admitted to acute care beds for whom continuous ECG data was available at the University of Virginia Medical Center. The 71 monitored beds are arranged in 3 units and are under the care of a variety of hospital services, principally Cardiovascular Medicine and Cardiothoracic Surgery. An institutional electronic data warehouse archived the electronic medical record (EMR) data, including admission, discharge, and transfer information. Patients with a length of stay of <72 hours were excluded from the analysis due to a lack of accumulated data to facilitate meaningful cumulative predictive analytics.

### EMR vital signs and laboratory results

At 15-minute increments, we recorded the most recent charted vital signs measurements and laboratory tests as described elsewhere.^15^ We excluded observations occurring after “Do Not Resuscitate” (DNR) or “Do Not Intubate” (DNI) orders or after transition to comfort measures-only (CMO).

### Cardiorespiratory dynamics measured from continuous ECG monitoring

#### Heart rate dynamics

We processed the continuous ECG with multiple QRS detection algorithms on the ECG lead with the highest signal to noise ratio. The three resulting heartbeat time series were combined to determine the probability density of each detected heartbeat. Low confidence beats were excluded from the analysis. We made observations every 15 minutes of the preceding 30 minutes and calculated the mean interbeat interval, the standard deviation or HR variability, and nonlinear dynamics of HR.^16-18^

#### ECG derived respiratory rate and respiratory sinus arrhythmia

We estimated the respiratory rate (RR) from both the cyclic variation in ECG waveform characteristics that result from respiratory movements and, when present, from the respiratory sinus arrhythmia peak in the frequency spectrum of heartbeat intervals. We analyzed 60-second interbeat interval time series windows containing 20 or more heartbeats. Windows overlapped by 75%. Details of how calculations were performed can be found in our previous publications.^19-22^

### Calculating the CoMET score

Details regarding how the CoMET score is calculated are exhaustively given elsewhere.^8, 9, 11, 13, 15^ Comprehensive clinical data that is incorporated into the CoMET score are shown in Table 1. Comprehensive clinical data that is incorporated into the CoMET score are shown in Table 1. The CoMET score is the fold-increase in probability of an event occurring in the next 8 hours, with a score of 1 meaning that the risk of the event occurring is the average risk for that particular patient in that particular unit. CoMET scores are calculated every 15 minutes to give a continuously updated estimation of the risk of imminent events. The event was emergent transfer to the ICU. The CoMET display as presented to clinicians is shown in Fig. 1 for 2 patients – a stable patient, and an unstable deteriorating patient.

**Table 1:** Data that is incorporated into the CoMET score

| Data Category | Examples |  |
| --- | --- | --- |
| Flowsheet vital signs | Heart rate |  |
|  | Respiratory rate |  |
|  | O2 saturation |  |
|  | Temperature |  |
|  | Blood pressure |  |
|  | Glasgow Coma Scale |  |
| Laboratory measurements | Basic and complete metabolic panels |  |
|  | Complete blood counts with and without diff |  |
|  | Arterial blood gases |  |
|  | Troponin levels |  |
|  | Blood coagulation tests |  |
| Continuous monitoring | Average and standard deviation of vitals | Heart rate |
|  |  | Respiratory rate |
|  |  | O2 saturation |
|  |  | Non-invasive blood pressure |
|  |  | Invasive blood pressure |
|  | ECG-derived parameters | Respiratory rate |
|  |  | Standard deviation of resp rate |
|  | Cross-correlation between signals | Heart rate and respiratory rate |
|  |  | Heart rate and O2 sats |
|  |  | Resp rate and O2 sats |
|  |  | Mean RR interval and ECG-derived resp rate |
|  | Inter-heart beat intervals parameters | Mean inter-beat interval |
|  |  | Standard deviation of RR intervals |
|  |  | Local dynamics score |
|  |  | Coefficient of sample entropy |
|  |  | Detrended fluctuation analysis |
|  |  | Probability of atrial fibrillation |

**Fig 1:**
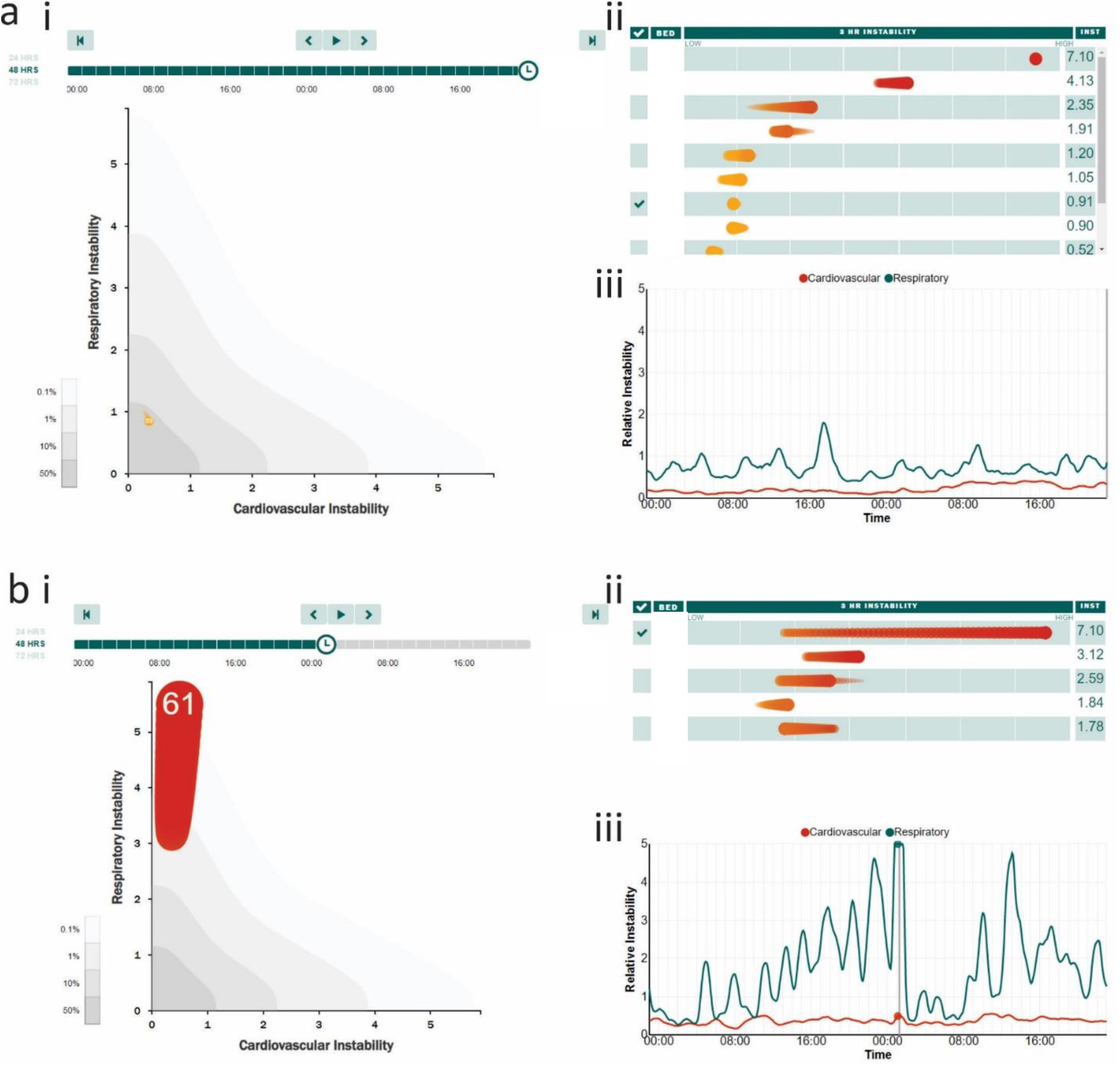
the CoMET display as presented to treating teams. Panel a shows 48 hour data from a patient in room 89, whose instantaneous cardiovascular and respiratory risk scores are low. Panel a,i shows the current CoMET for this patient (bed number is displayed in the head of the CoMET), which is small, and pale, reflecting the fact that the instantaneous risk is low. It is plotted on a graph with ‘cardiovascular instability’ on the x-axis, and ‘respiratory instability’ on the y-axis. The numbers are the fold increase in risk of an event within the next 6-12 hours. A score of 1 means that the risk of an event is at the average risk for that unit, with higher numbers meaning higher risk. The tail of the CoMET is barely visible, suggesting that the patient has been at this level of risk for the prior 3 hours. The contours on the graph delineate the expected percentile of patients in each contour of the graph, with darker shades of gray indicating more frequently populated areas – this patient is in the darkest gray area, and as such is exhibiting expected levels of risk for that unit – ‘nothing out of the ordinary’. Panel a,ii shows the ‘leaderboard’ – all of the patients on a given unit ordered from top to bottom based on their current CoMET score, highest to lowest. The 3-hour tail of the CoMET is also shown to demonstrate the degree of recent stability or instability. Individual patients can be selected for a deeper dive by checking the box to the left of their bed number (the check next to the patient in bed 89 can be seen). This brings up the graph shown in panel a,iii, which is a 24-, 48- or 72-hour graph showing the progress of the patient’s CoMET scores over the defined period, with cardiovascular instability shown as the red line, and respiratory instability shown as the green line. Panel b reflects the data from a different patient in bed 61, who appears much less stable. Their CoMET (b,i) is large and bright red, emphasizing the current high level of risk, mainly in respect of their risk of respiratory instability. The tail of the CoMET demonstrates that this instability has substantially progressed over the prior 3 hours, and that the patients CoMET now occupies a position on the graph which is rarely occupied (very pale contour), and ought to draw clinical attention to the patient if it has not done so already. The leaderboard in b,ii has this patient at the top (most unstable on the unit), along with the CoMET tail which is long and increasing, underscoring the increasing degree of risk and instability. Selecting this patient using the check box next to their bed number brings up the graph shown in panel b,iii, which demonstrates the evolution of CoMET over the prior 48 hours, showing recurrent and at times progressive respiratory instability in the face of relative stability of the cardiovascular risk. The instantaneous CoMET graph is deliberately selected at a time of peak respiratory instability (occurring at around 1 am on the day shown).

### Calculating cCoMET

Different to the CoMET score described above, which is an instantaneous estimate of risk of an imminent event, the cCoMET was developed to be a dynamic risk marker with memory. Calculation of the cCoMET was performed by comparing the instantaneous CoMET score to a dynamic baseline predicted risk, individualized for that patient. The baseline risk model used in the current study incorporated age, time since admission (minutes), admitting service, race, and sex. If the instantaneous CoMET score was below the baseline risk, the patient received a negative contribution to their cCoMET proportionate to the amount the CoMET was below the baseline predicted risk. If the patient’s CoMET scores remained below the baseline predicted risk, then their cCoMET would become progressively more negative over time. Conversely, if the instantaneous CoMET score was above the baseline predicted risk, the patient received a positive contribution to their cCoMET proportionate to the amount the CoMET was above the baseline predicted risk. If the patient remained above the baseline predicted risk, they would develop a progressively more positive cCoMET. Therefore, the final recorded cCoMET at the end of a patients stay reflects the aggregate degree of risk endured by the patient over that particular stay compared to the baseline model, and gives an idea of whether this was a ‘low risk hospitalization’ or a ‘high risk hospitalization’.

## Results

We retrospectively calculated the cCoMET scores of 8105 patients admitted to the acute care floor of the University of Virginia Medical Center from October 11, 2013 to September 1, 2015. After removal of patients whose stay was <72 hours, 5363 patients remained for analysis. The mean age of all patients was 64.6 years (range 18-90 yrs), while the mean length of stay was 8.5 days (range 3-122), with an average of 2.1 ICU days (range 0-104). 116 patients died (2.2%). When compared to the surviving patients, those patients who died were older (67.6 yrs vs 64.5), had longer lengths of stay (17.2 days vs 8.3), and averaged more days in the ICU (8.5 days vs 2.0) prior to their death. The vast majority of patients were classed as white (4362, 81%) on the electronic medical record, with significantly fewer being classed as black (897, 17%) or ‘other’ (104, 2%). Of the patients who died, 86 were white (74%), 27 were black (23%), and 3 were classed as ‘other’ (3%).

### cCoMET’s relationship to length of stay

Since cCoMET is cumulative over time, we examined whether there was a marked relationship between cCOMET and length of stay. If cCoMET increased monotically with length of stay, then it would not fit our purpose as an indicator of illness burden. However, Fig. 2 demonstrates there is no large dependence of cCoMET on length of stay. While the final cCoMET score was higher in those patients in the quintile of longest length of stay, and was progressively lower with each shorter length of stay quintile, these differences were small. LOS quintiles 1 thru 5 had median cCoMET scores of -0.44, -0.18, - 0.12, -0.12 and 0 respectively, (p=<0.001, Kruskal-Wallis test).

**Figure 2:**
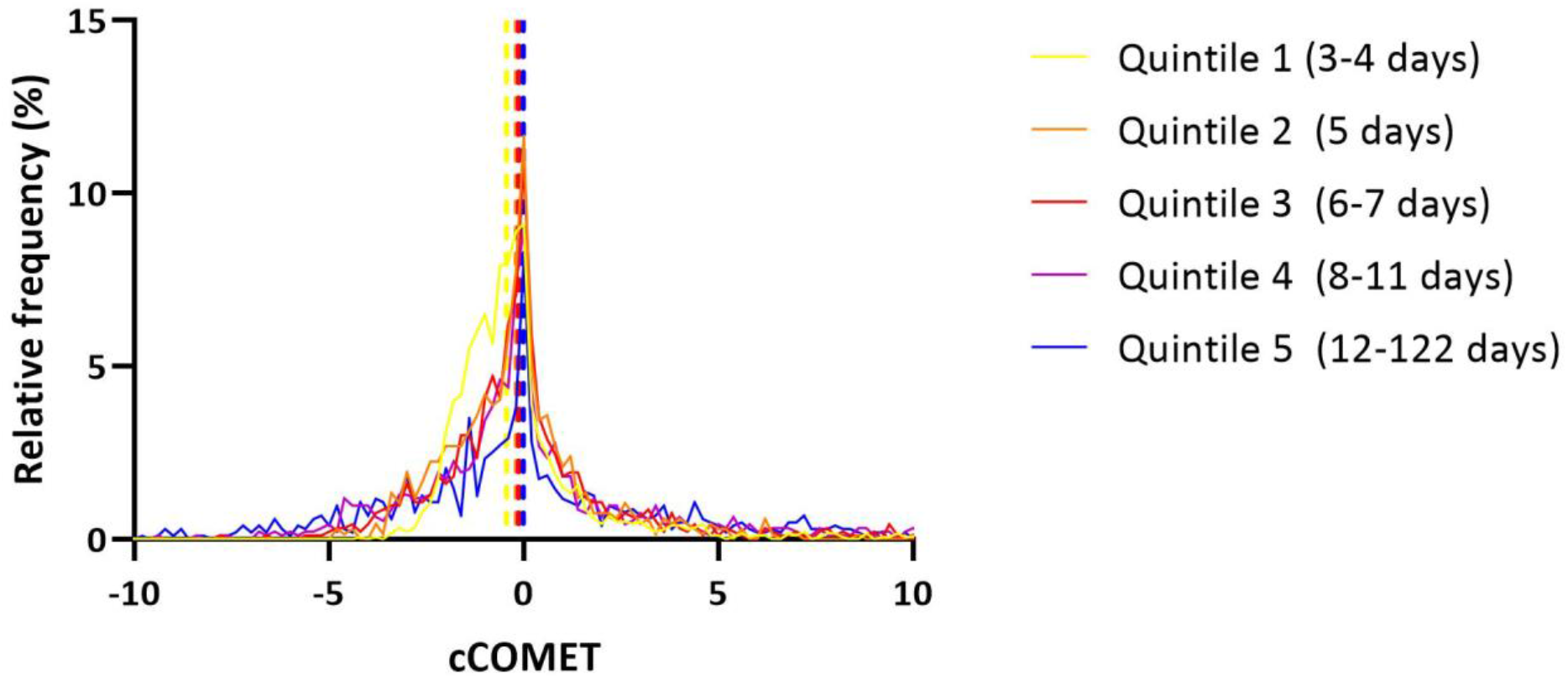
Relationship of cCoMET with length of stay. Dividing all patient up into quintiles based on their length of stay reveals that patients with higher lengths of stay have higher cCoMET scores (p<0.0001). Broken lines are median cCOMET scores in the different quintiles of lengths of stay. Inset uses a smaller x-axis to facilitate visualization of the differences between the groups.

### Distinct patterns of cCoMET evolution

Exemplar patterns of cCoMET evolution are shown in Fig 3. Panel a demonstrates the data from a patient who survived to discharge. Their CoMET score (green) was lower than the baseline model (blue) for the majority of their stay, and accordingly the cCoMET (red) became a progressively more negative value as time passed. This patient’s final cCoMET score was more negative than -5 on the day of discharge, suggesting that this was a ‘low risk hospitalization’. Panel b demonstrates another pattern in cCoMET score evolution – that of a patient who did *not* survive their hospitalization. In this patient, the CoMET score was persistently above the baseline model, and accordingly the cCoMET score became progressively more positive, until ultimately they died. At the point of death, the cCoMET was above 20.

**Figure 3:**
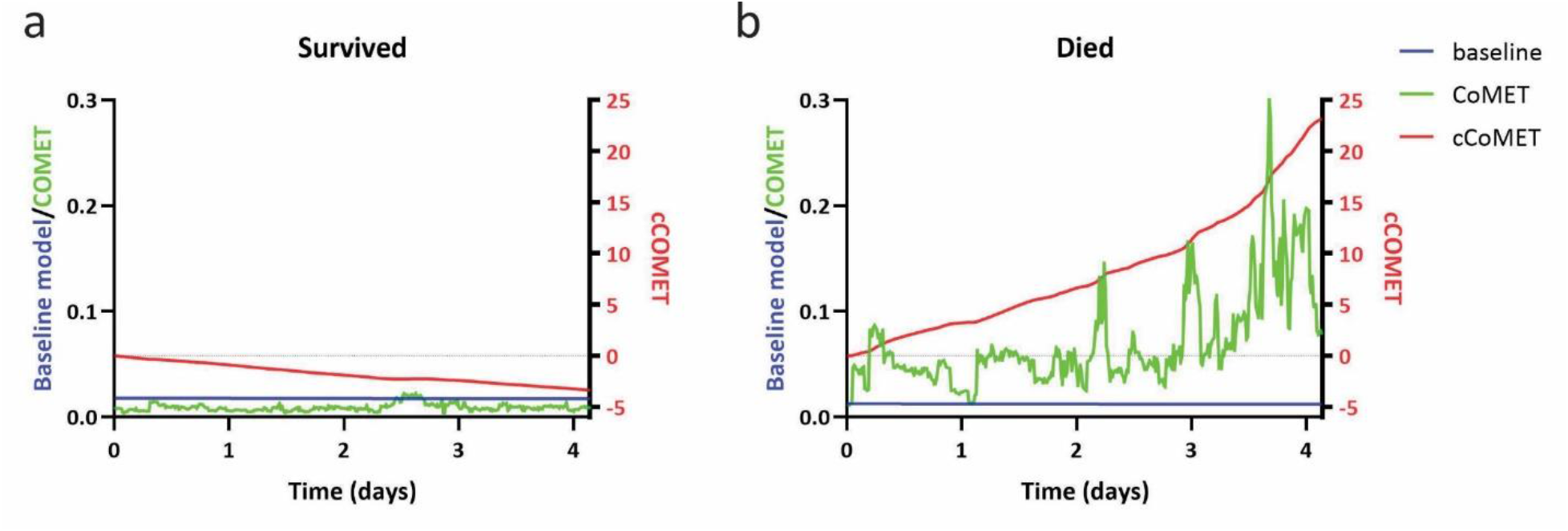
examples of cCoMET score evolution in two example patients. a. 4-day evolution of cCoMET score in a patient who survived hospitalization to discharge. Baseline risk is shown in blue. Actual CoMET score through admission is shown in green. cCoMET is shown in red. Because the CoMET score is persistently below the baseline predicted risk throughout the admission, the cCoMET score becomes progressively more negative than zero throughout the admission, being around -5 on the day of discharge. b. 4-day evolution of cCoMET score in a patient who died during this hospitalization. Colors are the same as in panel a. The CoMET score is persistently above the baseline predicted risk, and so cCoMET becomes progressively more positive through the admission, being around 23 just before death. x-axis numbers are probabilities of an event occurring in the selected CoMET model.

### Increasing cCoMET score was associated with mortality

Higher cCoMET scores were significantly associated with mortality. Fig. 4 demonstrates a rightward tail to the histogram in patients who died during this hospitalization, with a median cCOMET of 2.49 in patients who died versus -0.25 in those who survived (p<0.0001, Kolmogorov-Smirnov test).

**Figure 4:**
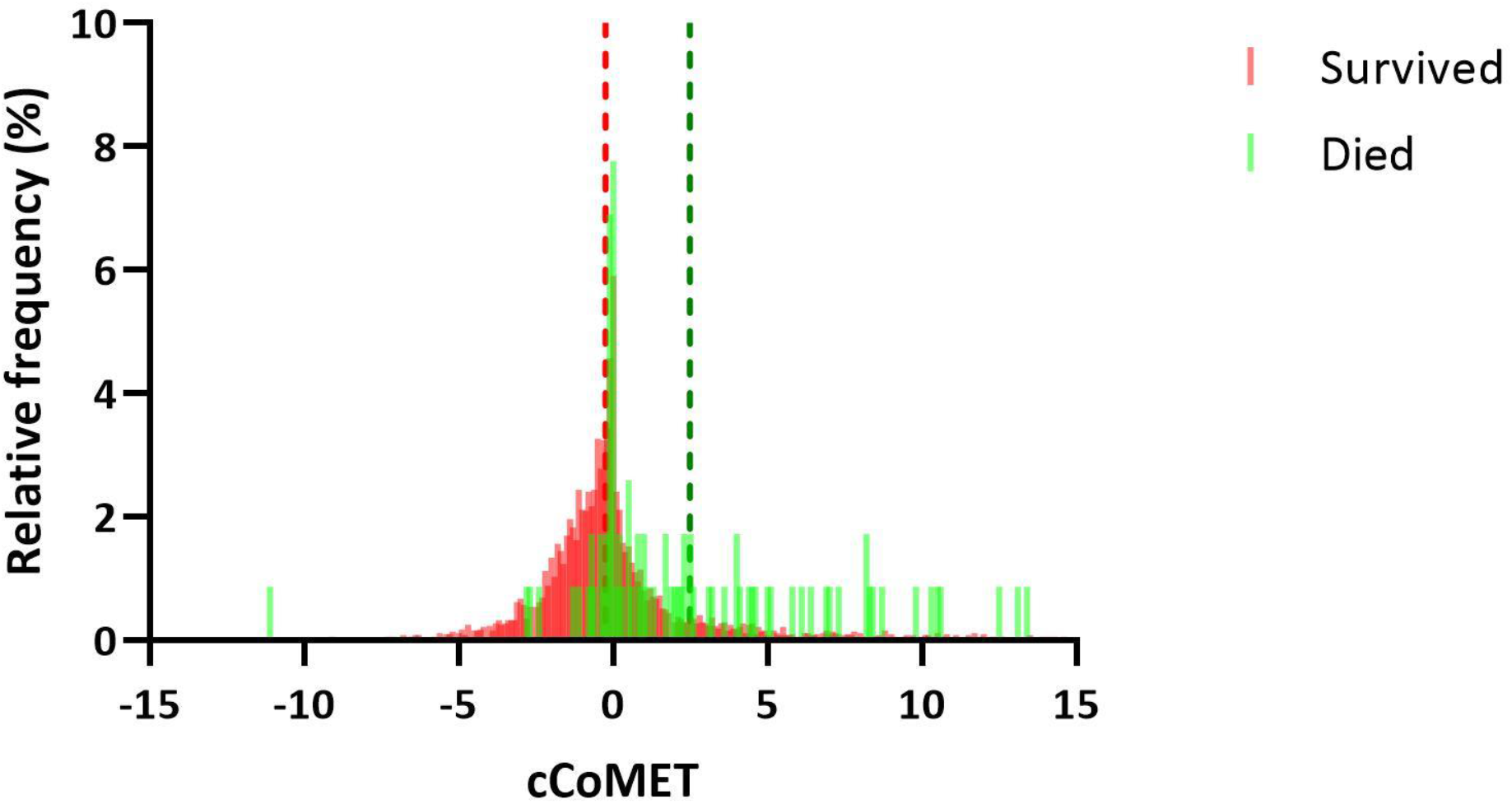
The relationship between cCoMET and mortality. a. Box-and-whisker plots of cCoMET score based on whether the patient survived (red dots) or died (green dots) during the index hospitalization. Mean cCoMET in surviving patients was 1.19, while it was 10.41 in those who died. b. histogram depicting the relative frequency of individual cCoMET scores by whether the patient survived (red bars) or died (green bars). The median cCoMET in each group is depicted by the broken line, and was significantly higher in those who died (2.49 vs -0.25, p<0.0001)

Fig. 5 demonstrates that in both patients who died (green circles) and those who survived (red circles), higher baseline CoMET scores were positively associated with higher final cCoMETs. Note that here the ‘baseline CoMET’ score is the initial CoMET score on admission minus the baseline risk estimate – negative values of this suggest a lower baseline risk compared to expected on admission, while positive values suggest a higher baseline risk compared to expected on admission.

**Figure 5:**
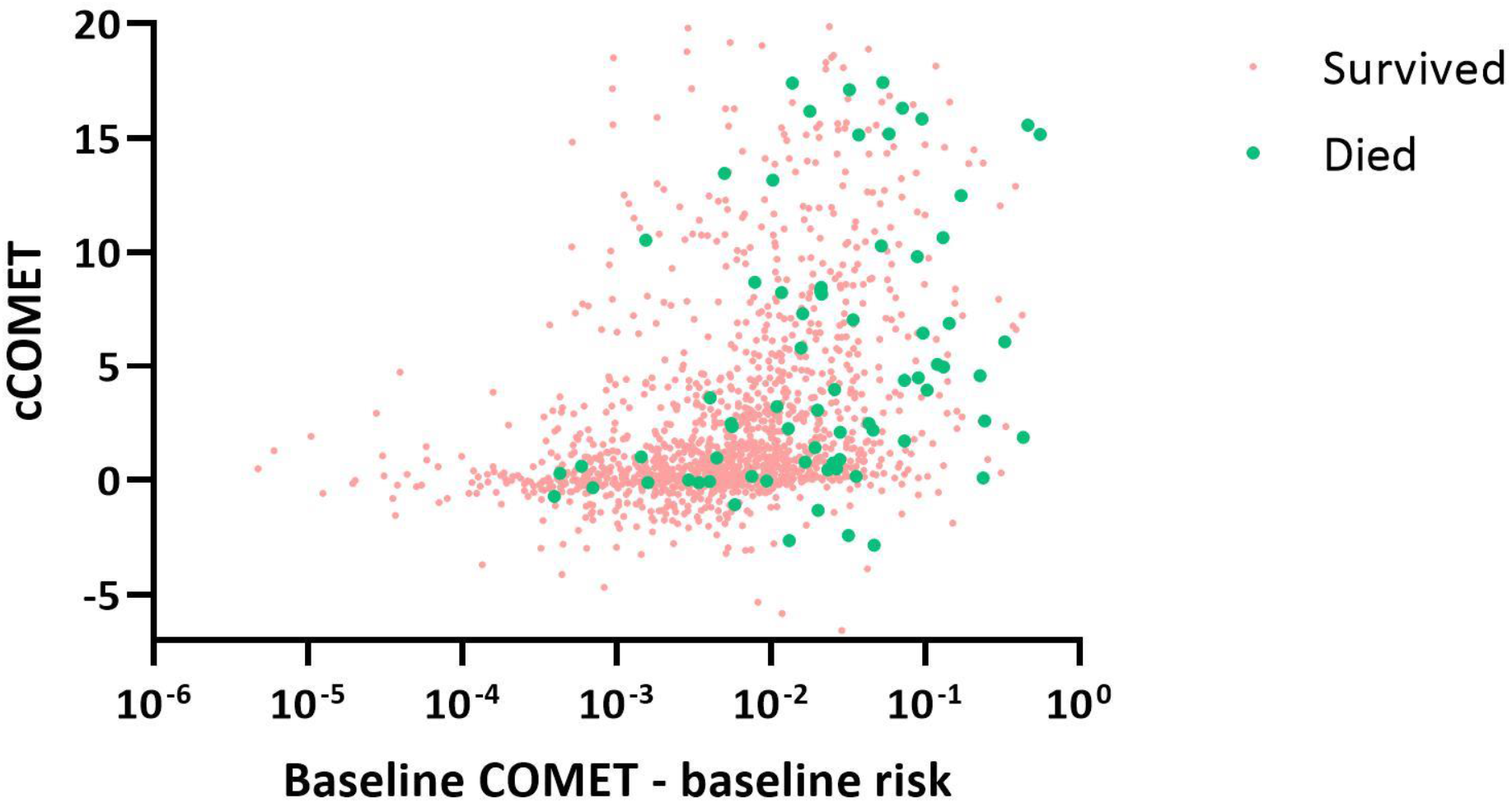
The relationship between baseline CoMET (the first CoMET score adjusted for by the baseline risk) and cCOMET in patients who survived (red dots) and those who died (green dots) during the index hospitalization. In both groups, higher baseline CoMET scores were associated with higher cCoMET scores.

### cCoMET adds information to the CoMET score when predicting mortality

Fig. 6 shows the log odds of death as a function of a logistic regression model using baseline factors (left panel), the cCoMET (central panel), and the last CoMET score reflecting the final 30 minutes of data (right panel) – the relationship between death and any of these parameters is steepest for the last CoMET prior to death (p <0.001), and as such this is the strongest predictor, while cCoMET is less effective (yet still has a significant p-value: 0.03) and baseline CoMET is the least effective (non-significant relationship: p = 0.10). One might speculate that the reason that last CoMET score outperforms cCoMET in this circumstance is that significant numbers of patients die from sudden and unpredictable illnesses that could not have been foreseen using a tool such as cCoMET. Such diagnoses include ventricular arrhythmia, aneurysmal rupture, intracranial hemorrhage, acute myocardial infarction, and pulmonary emboli.

**Figure 6:**
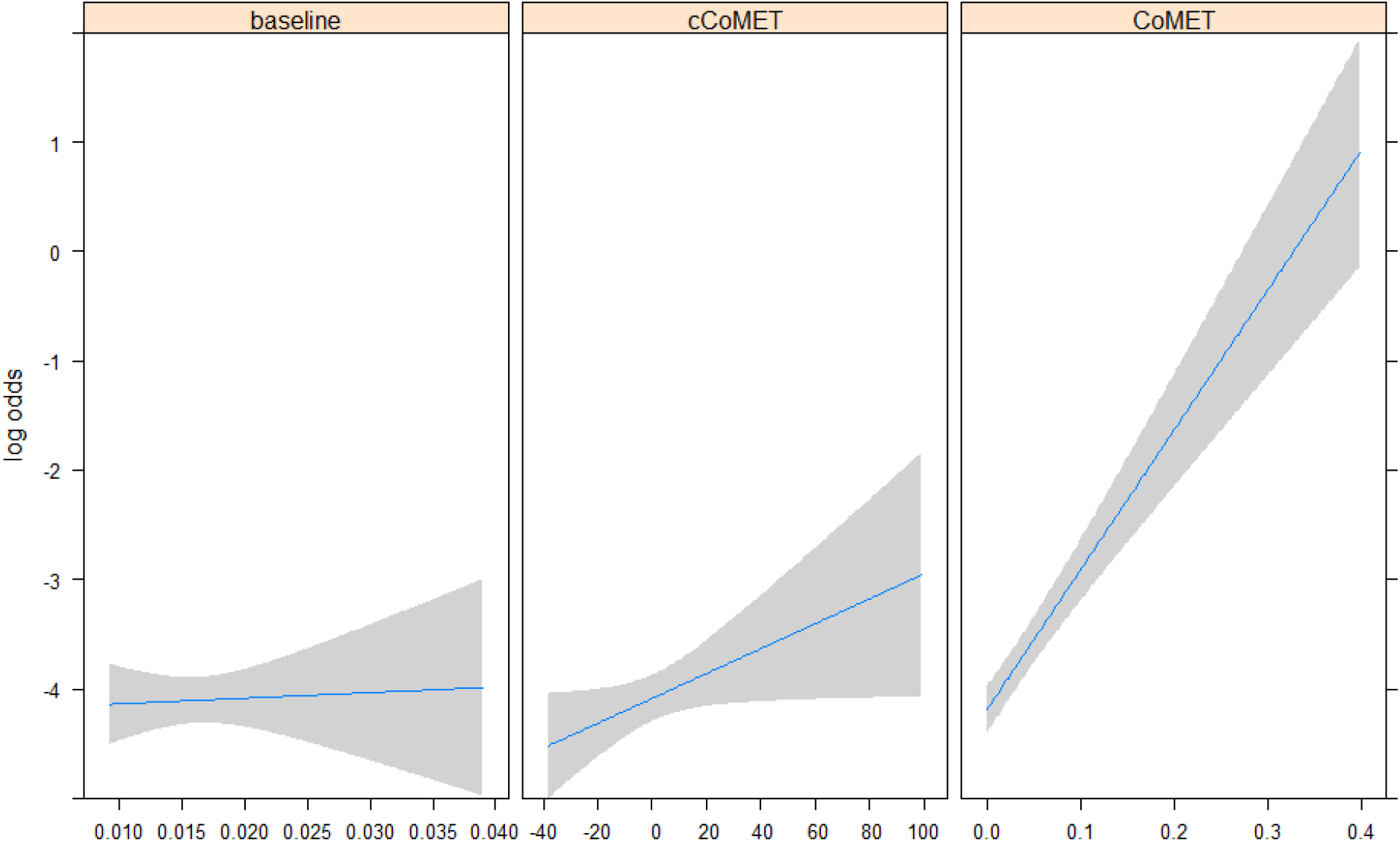
The log odds of death as a function of the outputs of logistic regression models using baseline factors only (left panel), the cCoMET score (central panel), and last CoMET score (right panel) – the relationship between death and any of these parameters is steepest for the last CoMET prior to death, and as such this is the strongest predictor, while cCoMET is less effective and baseline factors are the least effective.

### Sex differences in cCoMET

Fig. 7 is a histogram of final cCoMET by sex, demonstrating that males in the studied cohort are more likely to have higher (more positive) cCoMET scores, and hence to have high risk hospitalizations, than their female counterparts (median -0.16 vs -0.33, p<0.001 on Kolmogorov-Smirnov test).

**Figure 7:**
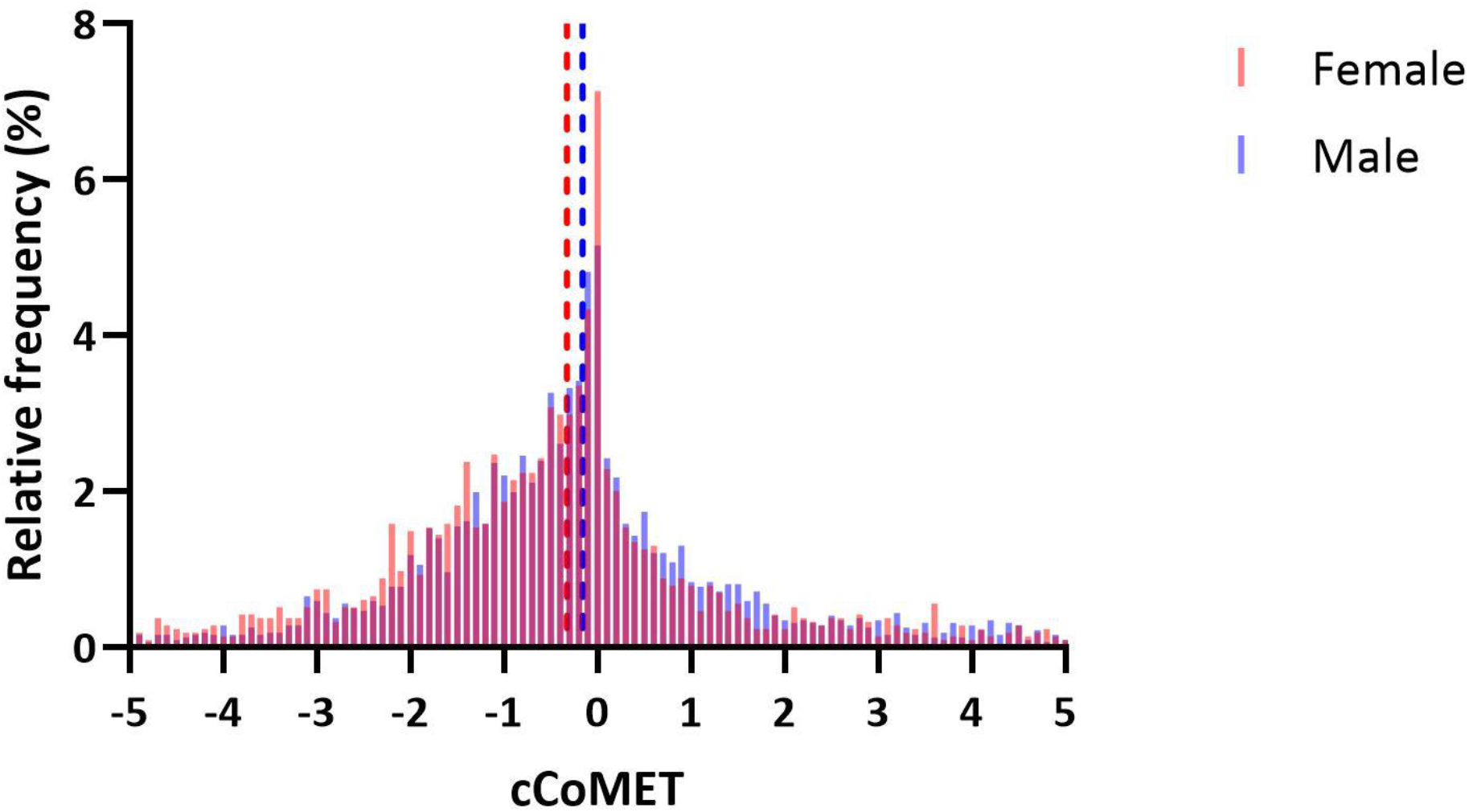
Sex differences in cCoMET score. Histogram depicts cCoMET score based on sex, showing that males (blue bars) have higher cCoMET scores than females (red bars, p<0.0001). Broken lines are median cCoMETs in the two groups.

## Discussion

The development of risk prediction models to assist clinicians evaluate high vs low risk hospitalizations, and weigh this information to avoid potentially preventable deteriorations and attain optimal outcomes in patients, is a developing and rapidly expanding field.^23^ The CoMET model referred to in this paper was developed from 146 patient years of vital sign and ECG waveform time series, encompassing 9232 ICU admissions, and 1206 clinician chart review identified episodes of unplanned intubation, sepsis or hemorrhage.^9^ The multivariate models that were developed from this data demonstrated C-statistics of 0.61-0.88 with respect to their ability to predict such events up to 24 hours prior to their occurrence, and give the clinician a dynamic mean fold-increase in the risk of that event occurring.

In this new work, we have studied how to use the CoMET score to produce a new metric, the cumulative CoMET, or cCoMET, score, by comparing each individual 15-minute CoMET score with respect to a baseline risk model, and then summing the results throughout the patients stay. The baseline risk model here was simplistic, comprising age, time since admission, admitting service, race, and sex. The blue lines on Fig. 3 show that the baseline risk model varied relatively little with time, while every-15-minute CoMET scores varied more markedly. CoMET scores above the baseline risk model added to the cCoMET score, while instantaneous CoMET scores below the baseline risk model subtracted from the cCoMET score. Thus, unlike the instantaneous CoMET score, cCoMET is a historical record of risk during a patient’s stay, and reflects not only on recent developments in the patients CoMET score compared to the baseline model, but on *all* CoMET scores since admission. One way to look at it is that the cCoMET reflects *accruing risk or indeed damage*, or lack thereof, occurring to a patient’s health status during a given hospital admission. A very positive cCoMET score suggests that a lot of damage was accrued to a patient’s health during a hospital admission, while a very negative cCoMET suggests that much less damage than expected occurred to a patient’s health during a hospital admission.

Our main findings are that cCoMET is crucially little affected by length of stay, and that higher (more positive) cCoMET scores portend a higher risk of dying in a given hospital admission. We have also found that a high baseline CoMET is related to an ultimately higher cCoMET in both patients who survived or died during the index hospital stay. Finally, cCoMET scores seems to be higher in males than females in our patient group. In performing this work, we have sought to extend the value of instantaneous risk scores, and to ensure that all valuable data accrued during a patient’s hospital stay is brought to bear when assessing their ultimate risk.

### Relationship to the work of others

The prediction of deterioration in hospitalized patients has traditionally utilized track-and-trigger systems, including the National Early Warning Score (NEWS) and others.^24-33^ These scoring systems have benefits including their relative ease of use and accessibility, yet suffer because they largely rely on vital signs and lab tests, which are variously intermittent, delayed, incorrect, unvalidated or indeed never taken.^34-37^ Few take account of crucial information contained within continuous cardiorespiratory monitoring. Potential benefits of the CoMET score are that it includes all of the above, and furthermore utilizes multiple models that have been trained on patients with specific causes of clinical deterioration identified by clinician review,^9^ and in doing so it is capable of learning the signatures preceding the development of specific clinical illnesses.^8^ However, the CoMET score is also an instantaneous score, based on vital signs, lab values and continuous cardiorespiratory monitoring parameters that are happening *now*.^13^ The novelty of the cCoMET score explored here is that it includes the ‘now’, but adds this to all of the predicted risk that has occurred *from admission to now*, to add vital information to current risk prediction. While one can argue that the instantaneous CoMET score (and the factors that it takes into account) has some degree of short term memory (i.e. lab values, vital signs and cardiorespiratory parameters do not vary with time at random, or stochastically, instead do bear some relation to the values that went before), it is not able to take account of all aspects of the risk since admission in as comprehensive a way that the cCoMET does. As such, the demonstrated relationships between cCoMET and mortality, baseline CoMET and gender are both novel and important, and emphasize the need for development of predictive analytics scores that incorporate ‘memory’, since, without this, important characteristics of the patient’s condition may be overlooked, negatively impacting on the performance of predictive analytics tools.

## Limitations

We omitted all patients with lengths of stay <72 hours, since it was felt that these patients did not have enough time to develop a meaningful cCoMET score, though this may have affected the data presented. The numbers of surviving patients was substantially more than the patients who died, limiting the analysis in this group, and limiting the statistical significance of the findings.

## Data Availability

All data produced in the present study are available upon reasonable request to the authors

## Notes

### Competing Interest Statement

The authors have declared no competing interest.

### Funding Statement

This study was funded in part by a philanthropic donation through the Frederick Thomas fund at the University of Virginia

### Author Declarations

Ethics committee/IRB of the University of Virginia gave ethical approval for this work

